# Towards Integrated Digital Health Systems for Nutrition and Food Security in Uganda: A Cross-Sectional Survey

**DOI:** 10.64898/2026.04.05.26350208

**Authors:** Amir A. Samnani, Nasser Kimbugwe, Elicana Nduhuura, Marriette Katarahweire, Benjamin Kanagwa, Katie Crowley, Audrey Tierney

**Affiliations:** School of Allied Health, University of Limerick, Limerick, Ireland; Health Research Institute, University of Limerick, Limerick, Ireland; Lero – the Research Ireland Centre for Software, University of Limerick, Ireland; School of Computing and Informatics Technology, Makerere University, Kampala, Uganda; Centre for One Health, University of Global Health Equity, Rwanda; Department of Computer Science & Information Systems, University of Limerick, Limerick, Ireland; Limerick Digital Cancer Research Centre, University of Limerick, Limerick, Ireland; Centre for Implementation Research, University of Limerick, Limerick, Ireland

## Abstract

Despite robust policy frameworks, Uganda’s digital health landscape is characterised by fragmentation—often termed “Pilotitis”—where stand-alone applications impede the integrated delivery of health, nutrition, and food security services. As part of the IGNITE project, this study mapped existing digital health systems (DHSs), identified systemic gaps, and explored opportunities and resource requirements for sustainable integration of existing Health, Nutrition and Food security data systems. The IGNITE project adopted a mixed-methods design; however, this paper reports findings from the first phase—a national cross-sectional survey conducted in Uganda. The survey mapped digital health, nutrition, and food security systems, identifying gaps, resource needs, and potential actions. Stakeholders from government, NGOs, academia, UN agencies, and frontline health workers were included using purposive and snowball sampling. Data were collected online and through field support. Of 134 respondents, 110 with ≥70% survey completion was included in the analysis. While 93% of respondents utilise digital tools (predominantly DHIS2 and mobile apps), only 20% reported full automated integration with national platforms. Critical barriers to interoperability included a lack of technical expertise (90%), insufficient DHIS2 training (82%), different data formats (77%), and infrastructure constraints (75%). Respondents identified workforce development (56%) and DHIS2 use and adoption (29%) as primary opportunities. Immediate priorities include staff training and provision of mobile hardware, while long-term strategies focus on standardised data formats (78%) and formalised governance frameworks for Integrated platforms (64%) and automated data exchange (56%). Uganda possesses a vibrant but disconnected digital ecosystem. Transitioning from isolated “data islands” to a cohesive system requires addressing the massive technical capacity gap and establishing mandated interoperability guidelines. The findings provide a data-driven roadmap for the Ministry of Health and partners to optimise digital health adoption, ensuring that nutrition and food security interventions are supported by a unified, evidence-informed digital architecture

**Author Summary:** Uganda uses many digital tools to track health and nutrition, but these systems often do not “talk” to one another. This creates “data islands” where important information about a child’s health or a family’s food security is trapped in one system and cannot be seen by other sectors. We wanted to map these systems to understand why they are fragmented and how to fix them. We surveyed 110 professionals, including government officials, international partners, and community health workers. We asked them what digital tools they use, what stops them from sharing data, and what resources they need to work more effectively. Most people use digital tools, but only 1 in 5 systems is fully connected to the national database. The biggest hurdles are a lack of technical skills, poor internet in rural areas, and different data formats that don’t match. Most workers feel they haven’t been trained well enough to manage these complex digital systems. To improve digital systems for health, nutrition and food security data in Uganda, we must move past small “pilot” projects and build one connected system. This requires urgent investment in training workers, providing tablets for data collection, and setting strict rules that require all new digital tools to be compatible with national systems.

## Introduction

Digital technologies are increasingly recognised as essential for strengthening health systems and improving population health, particularly in low- and middle-income countries (LMICs). The World Health Organization (WHO) defines Digital Health (DH) as “*the field of knowledge and practice associated with the development and use of digital technologies to improve health*” [1,2]. WHO and its Regional Office for Africa encourage African member states to adopt national digital health strategies aligned with the Global Strategy on Digital Health 2020– 2025 [3], emphasising interoperability, ethical data use, and equitable access. Regionally, the African Union Data Policy Framework guides data governance, while countries such as Kenya, Rwanda, and South Africa have established councils or interoperability frameworks to implement these commitments [3].

In Uganda, national policy increasingly emphasises digital transformation within the health sector. In 2021, the Ministry of Health (MoH) launched the Health Strategic Plan 2020/21– 2024/25 [4], aligned with the Third National Development Plan (NDP III) 2020/21–2024/25 [5] and Uganda Vision 2040 [6]. Building on this, in 2024 the MoH introduced the Health Information and Digital Health Strategic Plan 2020/21–2024/25 [7] to foster sustainable, ethically managed, and interoperable digital health solutions. These frameworks reflect Uganda’s commitment to a legal and regulatory environment that supports innovation, leading to the adoption of diverse digital health applications across the health system [3,8–10].

Despite these policy commitments, Uganda’s digital health ecosystem remains fragmented, with numerous stand-alone applications and information systems implemented by different stakeholders [2,7]. Many initiatives are developed independently using diverse technological architectures, resulting in limited interoperability and isolated “data islands” that restrict efficient information exchange across the health system [7]. This phenomenon, often termed “pilotitis,” has created information silos that impede seamless data sharing and communication [2,7]. Fragmentation is further exacerbated by limited standardisation, inadequate coordination among implementing partners, and poor integration with national health information platforms [7]. Consequently, the potential of digital health to improve outcomes and support integrated service delivery, including nutrition and food security interventions, remains under realised.

An illustrative example of the challenges posed by fragmented data systems in Uganda is observed in monitoring zero-dose and under-immunised children, as reported by the Uganda Learning Hub for Immunisation Equity [11]. The primary platform, District Health Information Software 2 (DHIS2), aggregates data for national reporting. While DHIS2 offers broad coverage and interoperability potential, it lacks individual-level data crucial for identifying zero-dose children [11]. Complementary platforms, such as the electronic Community Health Information System (eCHIS), capture household-level data with higher accuracy but remain pilot systems in selected districts and are not fully integrated into DHIS2 [11]. Other pilot systems, including Smart-Paper Technology, UgaVax, and EpiVac, vary in coverage, methodology, and reliance on partner support, producing inconsistent estimates and limiting longitudinal tracking across communities and facilities [11].

Challenges in data integration are also evident in nutrition information systems (NIS). Recent analyses of routine reporting platforms, such as DHIS2, highlight persistent gaps in standardisation, integration, and effective use of nutrition data [12]. Strengthening these systems requires adopting standardised nutrition indicators, optimising DHIS2 configuration, and enhancing analytics and dashboards to support data-driven decision-making [12]. The report also emphasises building technical capacity within Ministry of Health, improving data quality assurance, fostering cross-sectoral collaboration, and better integrating community-level nutrition data into national health information systems [12].

Uganda’s Nutrition Action Plan II (UNAP II) 2020/21–2024/25 provides a multisectoral framework to address malnutrition, aligning priorities with the National Development Plan III and engaging multiple ministries and agencies [13]. While UNAP II recognises the importance of strengthening nutrition information systems and the National Information Platform for Nutrition (NIPN), it does not clearly outline mechanisms for digital integration or interoperability across sectoral systems. Consequently, fragmentation may limit integrated reporting and analysis of nutrition and food security interventions.

In Uganda, the NIS consolidates nutrition indicators for facility- and district-level monitoring via DHIS2 [14], while NIPN uses multisectoral datasets for strategic policy analysis [15]. The Food Systems Dashboard, a global multi-institutional platform led by the Global Alliance for Improved Nutrition (GAIN), Johns Hopkins University, and partners, provides integrated data on food production, availability, diets, and livelihoods [16]. However, interoperability across these platforms remains limited, resulting in fragmented systems.

While these platforms provide important data, their fragmented nature highlights a critical evidence gap regarding how systems are used, connected, and governed in practice. Understanding this landscape is essential to identify opportunities for integration and inform the development of feasible, system-aligned solutions.

Considering this context, this paper presents findings from a mapping survey, a component of the broader mixed methods IGNITE project (Implementing DiGital Health SolutioNs for Food and NutrITion SEcurity in Uganda). The survey documents existing digital health systems, identified gaps and opportunities, assessed resource needs, and explores actions to promote more integrated and sustainable digital health, nutrition, and food security interventions in Uganda, providing evidence to guide coordinated, data-driven decision-making.

## Methods

### 2.1 Study design

This study is part of the broader IGNITE project, which utilised a mixed-methods design. This manuscript specifically presents findings from the cross-sectional survey (Phase I), which aimed to assess the current landscape of digital systems in place for recording and reporting health, nutrition, and food security data, including existing capacities, gaps, and resource requirements for implementing digital health solutions for food and nutrition security in Uganda.

### 2.2 Study participants

The survey targeted stakeholders engaged in health, nutrition, and food security programmes in Uganda, particularly those using digital tools, implementing digital solutions, or expressing interest in digitalisation. Participants included representatives from government institutions, United Nations agencies, academic institutions, non-governmental organisations (NGOs), and international non-governmental organisations (INGOs). Frontline health workers, including Village Health Teams (VHTs) and community health workers, were also included to capture operational perspectives from service delivery levels. The study sought to gather insights from actors involved in programme implementation, policy development, research, and digital system deployment related to health, nutrition, and food security

### 2.3 Study sites

The survey aimed to capture perspectives from stakeholders across Uganda. To complement online participation and ensure representation from frontline implementers, field-based data collection was conducted in three purposively selected urban and peri-urban regions: Kampala, Jinja, and Mbarara.

Site selection was guided by several considerations, including the presence of ongoing or recently implemented nutrition programmes within the past two to three years, representation across different regions of the country, varying levels of food insecurity, and diversity in urban and rural contexts. Kampala represents the national capital and a major innovation hub with relatively advanced digital systems and a concentration of national and international organisations. Jinja reflects an industrial and peri-urban setting characterised by mixed nutrition outcomes and moderate levels of digital adoption. Mbarara represents a rapidly urbanising district in south-western Uganda with a strong health system presence and active community engagement.

Within these regions, specific divisions, health sub-districts, and health facilities (Health Centre III and IV) were selected in consultation with relevant local authorities, including Chief Administrative Officers, District Health Officers, and City Health Officers.

### 2.4 Data collection methods

Data were collected using a structured survey designed on Qualtrics, to map existing digital health systems and identify gaps, opportunities, and resource needs related to digital health, nutrition, and food security systems in Uganda. The survey was administered both online and through facilitated completion by trained field teams.

The survey questionnaire consisted of six sections covering key domains relevant to digital health system integration. These included: (1) Respondent and organisational profile; (2) Mapping of current digital health systems (DHSs); (3) Understanding barriers and resource gaps; (4) Opportunities for improvement and potential solutions; (5) Planning for action; and (6) General feedback. In total, the questionnaire contained 23 questions, 16 of which were technical questions focused on digital systems, integration challenges, and implementation considerations.

The survey link was disseminated through multiple channels to reach a broad range of stakeholders. These included identified local actors, Ministry of Health representatives, and institutional partners such as Makerere University and Goal Uganda, the societal impact champion for the IGNITE project. The survey was also circulated through professional networks, including local WhatsApp groups related to health, nutrition, food security, and digital technologies, as well as through social media platforms such as LinkedIn.

In addition, the survey link was posted on the Emergency Nutrition Network (ENN) website to reach individuals and organisations working globally in nutrition, health systems, and food security. This platform was selected because it engages a wide network of practitioners, researchers, and organisations with relevant expertise in nutrition programming and systems strengthening.

To ensure inclusive participation, survey teams were mobilised in Kampala, Jinja, and Mbarara to facilitate responses from stakeholders who may have had limited digital literacy or lacked access to smartphones or internet connectivity. In such cases, trained field staff supported participants, including frontline health workers such as Village Health Teams and community health workers, in completing the survey questionnaire.

### 2.5 Sampling strategy and sample size

Participants were recruited using purposive sampling guided by a predefined stakeholder framework covering actors in digital health, nutrition, and food security systems in Uganda, including government, non-governmental organisations, implementing partners, and technical actors at national and sub-national levels.

Initial participants were identified through institutional partners (Makerere University and Goal Uganda) and relevant sectoral networks and platforms. Snowball sampling was then used to recruit additional eligible participants based on referrals from enrolled respondents

The study aimed for a target sample of 100 participants.

The study aimed for a target sample of 100 participants. No formal statistical sample size calculation was conducted, as the study was exploratory and mapping focused. Recruitment continued until sufficient representation from target groups were achieved.

### 2.6 Ethical considerations

The study was conducted in accordance with the principles of the Declaration of Helsinki. Ethical approvals were obtained from the Faculty of Education and Health Sciences Research Ethics Committee, University of Limerick, Ireland [2025_03_20_EHS] and the Research and Ethics Committee, Uganda [UNHL-2025-191]. Administrative approval to collect data from health facilities across the three regions was granted by the Director General of Health Services at the Uganda Ministry of Health. Subsequent site-level clearance to recruit and engage with participants was obtained from the respective District Health Officer, City Health Officers, and Hospital Directors.

Participation was voluntary, and all participants provided informed consent before completing the survey. For online participants, consent was obtained electronically via a consent form preceding the questionnaire. For field-based participants, consent was obtained verbally or in writing, as appropriate, with trained data collectors facilitating the process. Participants were assured that their responses would remain confidential and anonymised, and that data would be used solely for research and policy purposes.

### 2.7 Data analysis

Quantitative data from closed-ended survey questions were analysed descriptively using frequencies and percentages. Open-ended responses were analysed using thematic analysis, whereby two researchers independently coded the responses to identify recurring themes. Codes were reviewed and refined through consensus discussions to ensure consistency and reliability. Combining open and closed-ended questions allows participants to provide quick, structured data while also offering the freedom to explain complex local realities in their own words. For the researcher, this approach provides measurable statistics (the “what”) alongside rich, qualitative context (the “why”) to better understand systemic gaps, opportunities, and resource needs in digital health, nutrition, and food security in Uganda.

## Results

A total of 134 survey responses were received. Following a data quality audit, 24 (18%) were excluded for failing to meet the minimum 70% completion threshold. The final analytical sample comprised 110 respondents (82%), representing a robust cross-section of stakeholders engaged in Uganda’s health, nutrition, and food security sectors.

### 3.1 Respondent and Organisational Profile

The survey captured insights from 34 unique organisations. Government institutions constituted most of the sample (69%; n=76), reflecting their mandate in national service delivery and policy implementation. These were followed by International Non-Governmental Organisations (INGOs) (12%; n=13) and local NGOs (9%; n=10).

Work areas were predominantly concentrated in health (74%; n=81) and nutrition (61%; n=67). Notably, nearly half of the respondents (47%; n=52) were engaged in integrated health, nutrition, and food security services. Conversely, fewer participants reported primary involvement in food systems (15%; n=17) or digital systems (11%; n=12), suggesting a potential gap in specialised digital roles within these sectors.

Professionally, the sample was dominated by facility-based staff (41%; n=45), followed by office-based technical/support personnel (27%; n=30) and community-based frontline workers (22%; n=24). Policy-level actors (14%; n=15) and academic researchers (3%; n=3) represented the remainder of the cohort [Table 1].

**Table 1:** Organisational profile of survey respondents (n=110)

| Category and Variable | n | % |
| --- | --- | --- |
| **Q1. Type of Organisation** |  |  |
| Government | 76 | 69 |
| International NGO | 13 | 12 |
| Local NGO | 10 | 9 |
| Others | 8 | 7 |
| UN Agency | 3 | 3 |
| **Q2. Primary Area of Work (Select all that apply)** |  |  |
| Health | 81 | 74 |
| Nutrition | 67 | 61 |
| Integrated Services (Health, Nutrition & Food Security) | 52 | 47 |
| Food Security / Food Systems | 17 | 15 |
| Digital Systems | 12 | 11 |
| Others | 9 | 8 |
| **Q3. Primary Role in Organisation** |  |  |
| Facility-based staff | 45 | 41 |
| Office-based technical/support staff | 30 | 27 |
| Community-based frontline worker | 24 | 22 |
| Policy or decision-making level | 15 | 14 |
| Academic or research staff | 3 | 3 |
| Other | 3 | 3 |

### 3.2 Digital Systems Usage and Functionality

Of the 110 participants, 102 (93%) met the inclusion criteria for digital system usage (current or past three years). The digital landscape is characterised by the widespread adoption of DHIS2 (55%; n=56) and various mobile applications (39%; n=40). Electronic Medical Record (EMR) systems, including Open-MRS and Uganda-EMR, were utilised by 16% (n=16) of the sample. Specialist platforms for supply chain or workforce management were less prevalent.

In terms of utility, these systems primarily support health and nutrition service delivery (72%; n=73). Other significant functions include nutrition monitoring (45%; n=46), growth monitoring/counselling (38%; n=39), and client follow-up (28%; n=29). Digital applications for logistics, workforce management, and food security early warning systems (21%; n=21) showed lower levels of penetration.

Over half of the respondents (54%; n=55) reported that integration with the national DHIS2 platform is only partial, necessitating manual intervention. Furthermore, 25% (n=25) reported a total lack of integration, with only 20% (n=20) achieving fully automated data exchange

Consistent with this, respondents identified multiple barriers to integration, particularly lack of technical expertise (90%; n=92), insufficient training on DHIS2 integration (82%; n=84), and incompatible data formats across systems (77%; n=79). Infrastructure constraints (75%; n=77) and lack of interoperability (64%; n=65) were also widely reported, alongside governance and resource-related challenges.

Formal data-sharing mechanisms were limited, with only 35% (n=36) reporting established protocols, while others reported no agreements (25%; n=26), lack of awareness (16%; n=16), or ongoing processes (14%; n=14). Reflecting these gaps, most respondents reported limited internal technical expertise for interoperability (58%; n=59), with only 12% (n=12) indicating sufficient capacity [Table 2].

**Table 2:**
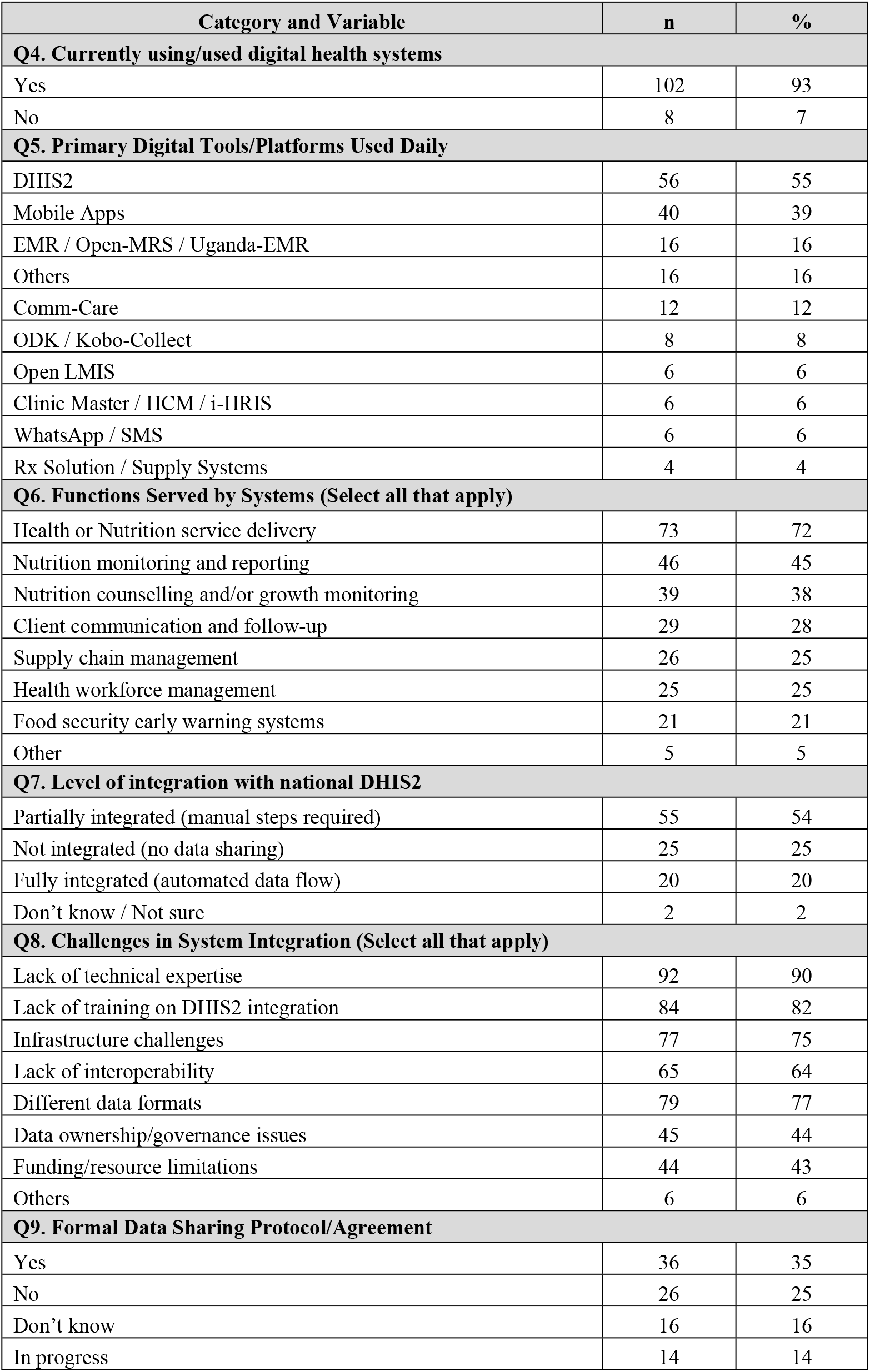
Digital system usage patterns and system functionality (n=102)

### 3.3 Understanding Barriers and Resource Gaps

A total of 102 respondents reported current or past use of digital health systems. In response to the question on the top challenges in using digital health systems for food security, nutrition, and health services, respondents highlighted a range of interconnected system and organisational-level barriers. The most prominent challenges related to training and capacity building (n=76), human resource constraints (n=68), and funding limitations (n=67), reflecting critical gaps in skills, staffing, and financial resources. These were closely followed by infrastructure and connectivity challenges (n=62), including unreliable internet access, limited devices, and power instability [Table 3].

**Table 3:** Reported barriers and resource constraints (n=102)

| Category and Variable | N | % |
| --- | --- | --- |
| **Q10. Internal Technical Expertise for Interoperability** |  |  |
| Yes, but limited | 59 | 58 |
| No | 23 | 23 |
| Don't know | 16 | 16 |
| Yes, sufficient | 12 | 12 |
| **Q11. Critical Resources Needed (Select up to 3)** |  |  |
| Training programmes for staff on DHIS2/Digital tools | 87 | 79 |
| Mobile devices / tablets for frontline collection | 81 | 74 |
| Stable internet connectivity for remote areas | 73 | 66 |
| Technical support team for troubleshooting | 37 | 34 |
| Standardised data guidelines | 24 | 22 |
| Pre-configured DHIS2 modules for Nutrition/FS | 20 | 18 |

Technical and system-level barriers were also widely reported, including system downtime, interoperability issues, and fragmented data systems (n=57), alongside broader data-related challenges such as lack of integration and continued reliance on manual processes (n=45). Governance and policy-related barriers, including low adoption, coordination challenges, and sustainability concerns (n=40), were noted less frequently but remain important [Table 4].

**Table 4:** Reported Challenges in the Use of Digital Health Systems across Food Security, Nutrition, and Health Services.

| Theme | Sub-theme | Quotes | Frequency (n) |
| --- | --- | --- | --- |
| **Training & Capacity Building (n=76)** | Training / Need for upskilling | "Training issues" / "Lack of training" / "Need training"/ "Refresher trainings needed" / "No refresher trainings" | 53 |
|  | Digital literacy | "Digital literacy gaps among staff/VHTs" / "Low skills on how to use digital reporting and data collection" | 15 |
|  | Technical expertise | "Technical expertise by end users, training" / "Staff unaware of alerts or digital notifications" | 8 |
| **Human Resources & Staffing (n=68)** | Staffing shortages | "Staffing shortages" / "Limited employees" / "Few implementing staff on ground" | 38 |
|  | Skill gaps / low IT competency | "Low IT/digital skills / skill gaps" / "Staff not trained for system use" | 20 |
|  | Personnel availability | "No health information assistant / insufficient personnel" | 10 |
| **Funding & Resource Constraints (n=67)** | Insufficient funds | "Funding insufficient / lack of funds" / "Insufficient funds to facilitate the process" | 32 |
|  | Limited resources / devices | "Limited resources / devices / tablets / computers" / "Shortage of tablets, computers, iPads" | 25 |
|  | Short funding cycles / delays | "Short funding cycles / delayed resources" / "Funding gaps, delayed system updates" | 10 |
| **Internet & Connectivity Challenges (n=62)** | Connectivity issues | "Internet access issues / network problems" / "Poor network connectivity" | 37 |
|  | Slow / unstable internet | "Slow internet / unstable / data bundles expensive" / "Internet bundles too expensive" | 15 |
|  | Rural / remote connectivity | "Connectivity in rural areas" / "Internet access in remote areas" | 10 |
| **Technical & System Challenges (n=57)** | System breakdown / downtime | "System breakdown / downtime / errors" / "System downtime during critical times" | 25 |
|  | Interoperability / integration | "Interoperability / integration issues" / "Lack of interoperability between ODK and DHIS2" | 20 |
|  | Platform / offline limitations | "Multiple platforms not syncing / offline limitations" / "Limited offline capabilities of digital tools" | 12 |
| **Infrastructure & Equipment Challenges (n=60)** | Limited devices / computers | "Limited computers / tablets / devices" / "No computers or tablets" | 30 |
|  | Power / electricity issues | "Power outages / power instability / no backup" / "Frequent power outages" / "Power backup lacking" | 20 |
|  | Lack of infrastructure | "No equipment / infrastructure lacking" / "No infrastructure, no tools to use" | 10 |
| **Data & Interoperability Challenges (n=45)** | Data fragmentation | "Data fragmentation / multiple formats" / "Different data formats" | 20 |
|  | Lack of integration | "Lack of integration of nutrition/health data" / "Data not integrated across ministries" | 15 |
|  | Manual entry / incomplete systems | "Manual entry still needed / data not in systems" / "Data still being manually entered into registers" | 10 |
| **Governance, Policy & Cultural Barriers (n=40)** | Low adoption / reluctance | "Low adoption / reluctance to adopt systems" / "Uptake of digital health systems by Ministry of Health is low" | 15 |
|  | Ministry coordination / sustainability | "Ministry coordination / sustainability issues" / "Sustainability and ownership issues" | 10 |
|  | Cultural / language adaptation | "Need language/cultural adaptation" / "Solutions should accommodate local languages and cultural contexts" | 5 |

Consistent with these findings, most respondents reported limited internal technical expertise for interoperability (58%; n=59), with only 12% (n=12) indicating sufficient capacity.

In terms of priority resource needs, respondents most frequently identified training programmes for staff (79%; n=87), provision of mobile devices for frontline data collection (74%; n=81), and improved internet connectivity (66%; n=73) as critical enablers for strengthening digital system use.

### 3.4 Opportunities and Solutions

Respondents (n=110) highlighted key opportunities aligned with health system building blocks [Table 5]. For workforce development (n = 109), continued training and capacity building was prioritised by 61 respondents (56%), digital literacy by 14 (13%), mentorship by 11 (10%), IT personnel availability by 17 (16%), and community health worker engagement by 6 (6%). To improve digital health information systems (n = 91), deepening DHIS2 adoption (n = 32; 29%), data collection and quality improvement (n = 21; 19%), EMR linkages/interoperability (n = 19; 17%), and dashboard enhancements (n = 7; 6%) were highlighted. Leadership and governance opportunities (n = 90) included NGO/partner coordination (n = 28; 26%), rollout of new guidelines (n = 24; 22%), and eHealth strategy implementation (n = 18; 16%). Financial opportunities (n = 50) focused on donor funding (n = 25; 23%) and government budget allocations (n = 16; 15%). Service delivery improvements emphasised system integration (n = 32; 29%) and mHealth adoption (n = 8; 7%), while access to technology prioritized computers (n = 16; 15%), tablets (n = 10; 9%), ICT equipment (n = 8; 7%), and power supply (n = 5; 5%).

**Table 5:**
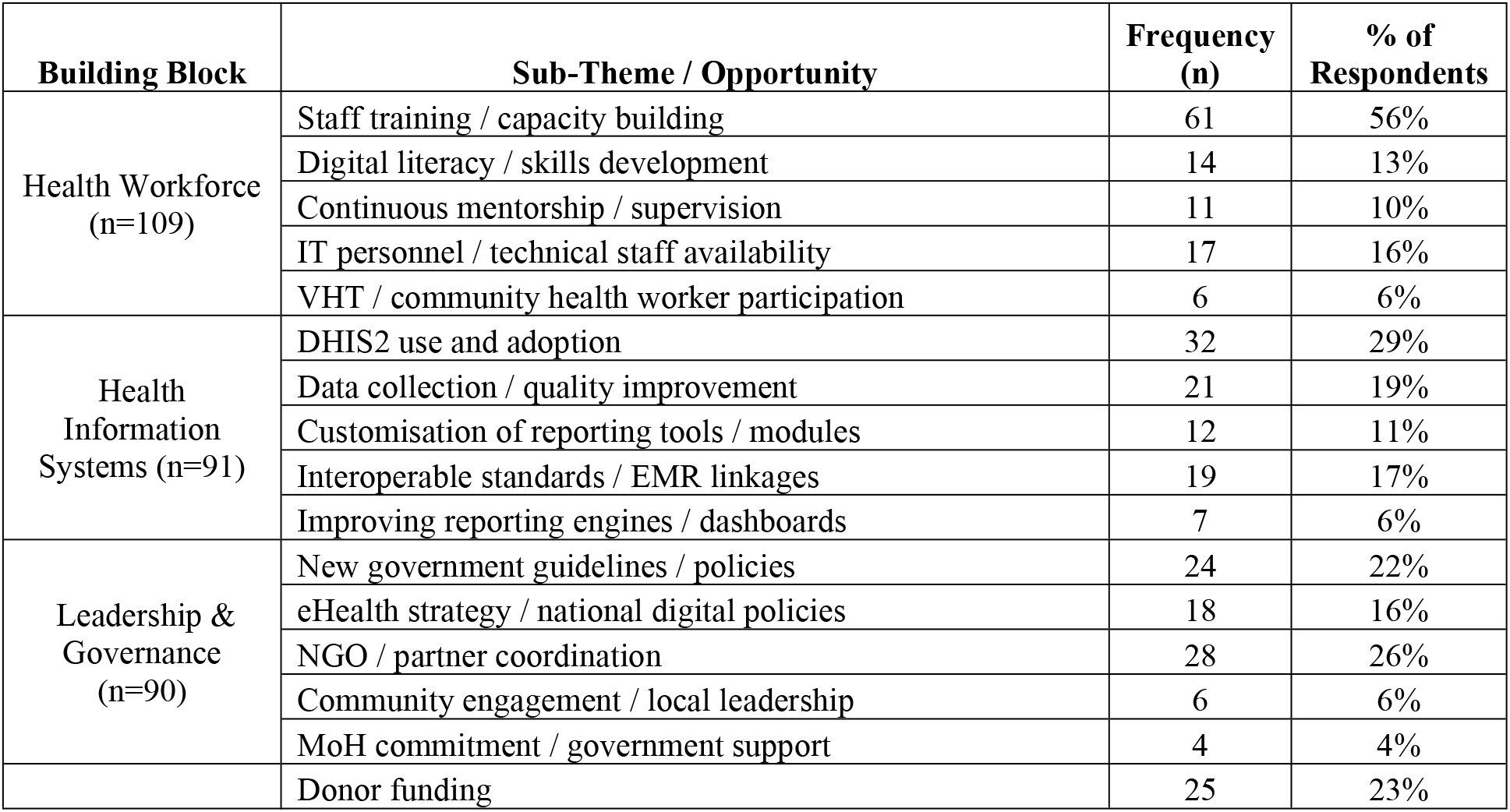
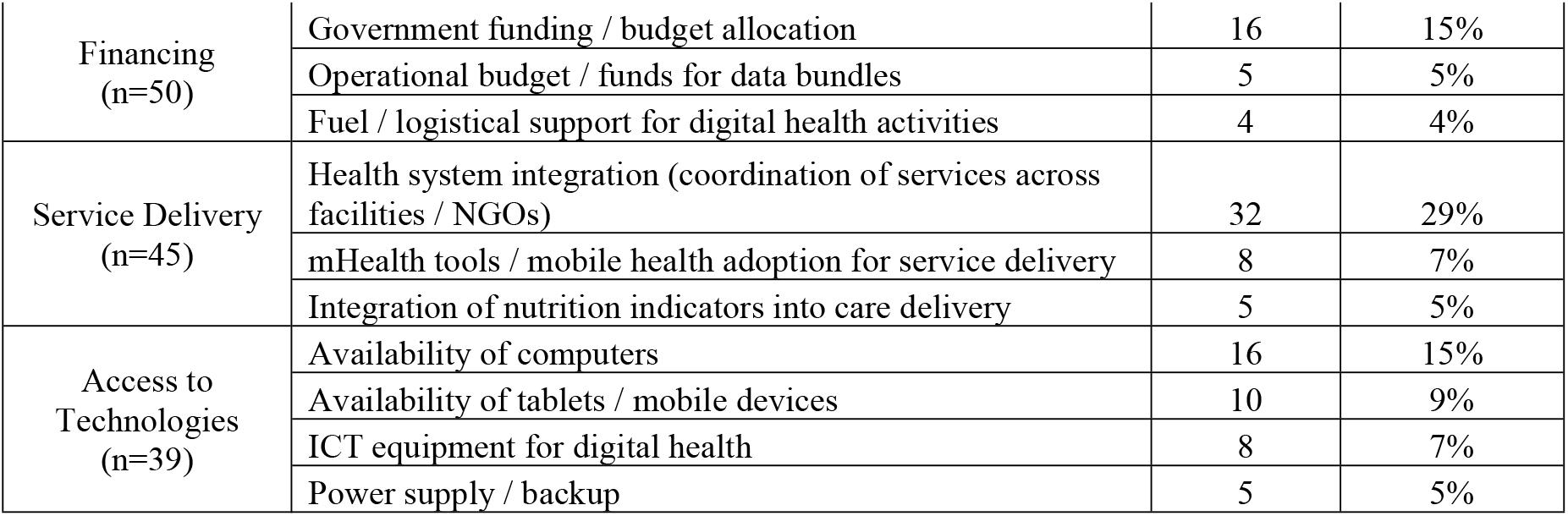
Perceived Opportunities to Strengthen Digital Health Systems for Nutrition, Food Security, and Health Services.

Preferred solutions for integration included training frontline workers (59%), public–private partnerships for infrastructure/connectivity (52%), mobile apps with offline DHIS2 sync (37%), pre-configured DHIS2 modules (36%), cloud-based management systems (36%), mandated DHIS2 compatibility (35%), and interoperability guidelines/taskforce (23%).

### 3.5 Planning for Action

Stakeholders recommended integrating existing digital health tools, identifying a diverse landscape of mobile-based platforms (n=38), such as ODK/KoboCollect and mTrac, alongside server-based systems (n=19) including HRIS and eLMIS. The inclusion of referral tracking (n=13), decision-support dashboards (n=7), and electronic medical records (n=5) underscores a critical demand for end-to-end integration across clinical, logistical, and governance functions.

To strengthen system functionality, respondents prioritised four key technical enhancements: standardised data formats (66%; n=73), user-friendly dashboards (58%; n=64), comprehensive training (55%; n=61), and automated data-sharing mechanisms (55%; n=60) [Table 6].

**Table 6:** Action priorities for digital system improvement and sustainability (n=110)

| Category and Variable | N | % |
| --- | --- | --- |
| **Q13. Technical or Process Improvement Options (Select 3)** |  |  |
| Standardised data formats | 73 | 66 |
| User-friendly dashboards | 64 | 58 |
| Better training on existing tools | 61 | 55 |
| Automated data sharing tools | 60 | 55 |
| Shared infrastructure | 27 | 25 |
| Other | 0 | 0 |
| **Q14. Long-term Sustainability Strategies (Select up to 3)** |  |  |
| Standardised data formats | 86 | 78 |
| Integrated platforms | 70 | 64 |
| Automated data exchange tools | 62 | 56 |
| Mobile data collection | 50 | 45 |
| Shared infrastructure | 27 | 25 |

Immediate priority actions to bridge current gaps centred on capacity building for frontline users (37%; n=41) and the reinforcement of digital infrastructure (30%; n=33), specifically regarding hardware access and reliable connectivity. Furthermore, increased resource mobilisation (21%; n=23) was deemed essential for operational sustainability. Secondary measures included addressing human resource shortages and improving multi-stakeholder coordination [Table 7].

**Table 7:** Immediate Actions for Strengthening Digital Health Systems.

| Theme | Sub-theme | Verbatim Response | Freq |
| --- | --- | --- | --- |
| **Capacity Building & Training (n=41)** | **Staff Training & Sensitisation (n=24)** | Sensitisation of all health workers | 3 |
|  |  | Training staff on digital health systems | 5 |
|  |  | Training of all staff on digital systems | 4 |
|  |  | Training staff on tools to use | 4 |
|  |  | Train frontline health workers, providing fast internet and keeping the system available for access every day | 3 |
|  |  | Train staff about digital health systems | 3 |
|  |  | Train frontline staff on the usage and adoption | 2 |
|  | **Continuous Learning & Mentorship (n=6)** | Continuous trainings and educating the community | 2 |
|  |  | Organise refresher training and mentorship sessions for health educators and VHTs | 2 |
|  |  | Conduct training and mentorship for staff on data entry and analysis | 2 |
|  | **Orientation & Skills Development (n=7)** | Immediate training about digital health system; Orientation of HMIS | 2 |
|  |  | Conduct CMEs on digital data management by IT personnel to all staff | 1 |
|  |  | Capacity building trainings and seminars | 2 |
|  |  | Ensure continuous capacity building of staff due to turnover rates | 2 |
|  | **Digital Literacy (n=4)** | Training on DHIS2 | 2 |
|  |  | Staff training on digital data management | 2 |
| **Funding & Resource Mobilisation (n=23)** | **Financial Support (n=15)** | Fundraising | 2 |
|  |  | Increase on funding the process and rollout | 2 |
|  |  | Mobilize for fundings to coordinate the systems | 2 |
|  |  | Funding digital health systems | 2 |
|  |  | Increased funding for integrated digital health systems | 3 |
|  |  | Funding to train more staff | 2 |
|  |  | More funding to health facilities | 2 |
|  | **Resource Allocation & Sustainability (n=6)** | Funds to facilitate trainings and procure laptops, procure data bundles | 2 |
|  |  | Funds to help in consistent workflows/operations like support trainings, procure computers, fuel for generator etc | 2 |
|  |  | Funding to acquire enough computers and backups | 2 |
|  | **Partnerships & Donor Engagement (n=2)** | Collaborating with governments, donors, and private sector entities to support funding and resource mobilisation | 1 |
|  |  | Further soliciting for donor funding to ensure effective and efficient utilization of digital health system | 1 |
| **Digital Infrastructure & Connectivity (n=33)** | **Hardware & Devices (n=9)** | Availing electronic devices to staff | 2 |
|  |  | Provide tablets to us for real-time data entry | 2 |
|  |  | Providing laptops and tablets to all staff | 2 |
|  |  | Buy computers and install apps | 1 |
|  |  | Provide enough mobile devices (tablets/smartphones) | 2 |
|  | **Internet & Power Connectivity (n=18)** | All time internet connection | 3 |
|  |  | Ensuring the internet is available | 3 |
|  |  | Provision of Internet connectivity | 2 |
|  |  | Stable Internet connectivity | 2 |
|  |  | Extend reliable power (solar system) | 2 |
|  |  | Install power, internet | 2 |
|  |  | Improve internet coverage | 2 |
|  |  | Stable power sources | 2 |
|  | **System Infrastructure & Integration (n=6)** | Development of infrastructure for digital health systems and employment of skilled workforce | 2 |
|  |  | Developing infrastructure to integrate use of digital tools to a common dashboard | 2 |
|  |  | Digitalisation of all records | 1 |
|  |  | Improvement in infrastructure development | 1 |
| **Human Resources & Staffing (n=9)** | **Staffing Gaps & Recruitment** | To have technical staff i.e. IT and Data managers | 2 |
|  |  | Recruitment of Information Technologists, Data Officers or Health informatics officers | 2 |
|  |  | Increase number of staff | 2 |
|  |  | More staff needed | 2 |
|  |  | To increase human resource | 1 |
| **Coordination, Governance & System Strengthening (n=11)** | **Coordination Mechanisms (n=5)** | Coordination between different stakeholders/departments | 2 |
|  |  | Government-NGO partnerships | 2 |
|  |  | Physical meetings with relevant stakeholders | 1 |
|  | **Policy & Leadership (n=3)** | No Unique identifier policy; Government leadership on all digitization efforts | 2 |
|  |  | Support guideline development for digitization | 1 |
|  | **Institutional Strengthening (n=3)** | Developing institutional capacity to facilitate digital health capabilities | 2 |
|  |  | Partnerships and collaboration with other organisations and government entities | 1 |
| **Community Engagement & Service Delivery (n=7)** | **Frontline & Community Empowerment (n=7)** | Community health sensitisation | 2 |
|  |  | Continuous trainings and educating the community | 2 |
|  |  | Equip VHTs and community health educators with smartphones/tablets | 2 |
|  |  | Improve internet connectivity for community health workers | 1 |

## Discussion

This national mapping survey offers a multi-level perspective on the digital health, nutrition, and food security landscape in Uganda. By engaging a diverse cohort of 34 organisations, ranging from system level actors to frontline community health workers,the study provides a comprehensive overview of the operational realities and systemic challenges inherent in digital transformation.

Our findings reveal a significant “adoption-integration paradox”; while digital health platforms such as DHIS2, mobile applications, and Electronic Medical Records (EMRs) are ubiquitous (93% usage), their utility is severely hampered by limited systemic cohesion. Only 20% of respondents reported full automated integration with national platforms, leaving the majority reliant on inefficient, error-prone manual data entry. This “data island” phenomenon reflects a persistent state of fragmentation previously documented by Ddamba et al. (2025), who noted minimal interoperability in public EMR systems, particularly across pharmacy and commodity modules [17]. Furthermore, Kiwanuka et al. (2026) observed that widespread DHIS2 deployment has not translated into real-time program planning due to these operational silos [18]. Our results corroborate these findings, suggesting that the “pilotitis” observed in the Ugandan health sector is a structural rather than a technological failure, where the proliferation of tools has outpaced the development of integrative architecture.

The identified challenges highlight a profound “human-infrastructure gap.” The overwhelming lack of technical expertise (87%) and insufficient DHIS2 integration training (79%) suggest that digital health investments have historically prioritised software procurement over human capital development. These findings align with recent district-level performance analyses, which identified human resource shortages and system errors as primary deterrents to digital adoption [18].

However, this study adds critical nuance by quantifying broader systemic impediments, such as funding constraints (61%) and governance barriers (36%), which were less visible in previous localized studies. The persistence of “fragmented data formats” (75%) points to a lack of shared technical standards at the central level. This suggests that without a mandated, unified data dictionary and governance framework, digital tools will continue to operate as isolated silos, failing to provide the multisectoral insights necessary for addressing complex issues like food security and malnutrition.

The alignment between identified resource needs, specifically mobile hardware, stable connectivity, and technical support and the proposed long-term measures indicates a clear strategic path forward. Respondents effectively outlined a two-tiered roadmap: immediate operational stabilisation through hardware and training, followed by structural reform via standardised data formats and automated exchange protocols. These priorities echo global best practices for digital health in resource-constrained settings, emphasising that interoperability is contingent upon “contextualised technical standards” and robust “institutional governance.”

The identified need for pre-configured DHIS2 modules specifically for nutrition and food security is particularly significant. It suggests that the current national architecture is not yet fully optimised for multisectoral SDG monitoring, necessitating a shift from “general health” reporting to “integrated service” surveillance.

A primary strength of this study is its breadth, capturing a snapshot of 34 diverse organisations within Uganda that could be representative of the national landscape. Nevertheless, the heavy concentration of respondents within the health sector limits the generalisability of findings across other critical sectors like Agriculture or Social Protection. To mitigate this, planned consultative workshops with the Uganda Bureau of Statistics and relevant multi-sectoral ministries will be essential. This cross-sectoral validation will ensure that the resulting roadmap for the IGNITE project is not only technically sound but also operationally viable across the entire food security and nutrition ecosystem.

In conclusion, while Uganda has achieved high levels of digital tool adoption, the lack of seamless interoperability remains the primary bottleneck to data-driven governance. To move beyond fragmented “pilot” initiatives, three strategic pillars must be prioritised:

1. Institutionalised Capacity Building: Moving from one-off trainings to integrated digital literacy for all health cadres.
2. Infrastructural Resilience: Ensuring that the “digital divide” is bridged through stable connectivity and hardware access for frontline workers.
3. Mandated Interoperability: Enforcing standardised data protocols and formal governance structures to enable a unified, scalable ecosystem.

Addressing these structural requirements is critical to transforming Uganda’s digital health landscape into a powerful instrument for achieving sustainable health, nutrition, and food security outcomes.

## Data Availability

All relevant data are within the manuscript and supporting files

## Author contributions

Funding acquisition: Audrey Tierney with Research Ireland

Conceptualisation: Amir A. Samnani, Katie Crowley, Audrey Tierney

Data collection: Amir A. Samnani, Nasser Kimbugwe, Elicana Nduhuura

Data Analysis: Amir A. Samnani

Project administration: Amir A. Samnani, Nasser Kimbugwe, Elicana Nduhuura, Katie Crowley, Benjamin Kanagwa, Audrey Tierney

Writing – original draft: Amir A. Samnani

Writing – review & editing: Nasser Kimbugwe, Katie Crowley, Benjamin Kanagwa, Audrey Tierney

## Acknowledgements

The authors would like to thank Abey Jose and Edith Bannerman for their valuable contributions to the broader IGNITE project activities. While not directly involved in the preparation of this manuscript, their efforts in supporting project implementation, coordination, and delivery of key project outputs greatly facilitated the work reported here

## Data availability statement

All relevant data are within the paper

## References

1. Global strategy on digital health 2020-2025. Geneva: World Health Organization; 2021. Licence: CC BY-NC-SA 3.0 IGO. https://iris.who.int/server/api/core/bitstreams/1f4d4a08-b20d-4c36-9148-a59429ac3477/content [last accessed: 23/03/2026]

2. Wamema, J., Amiyo, M., & Nabukenya, J. (2025). Standardising digital health interventions in Uganda’s health system using an enterprise architecture approach. BMC Digital Health, 3(1), 73. 10.1186/s44247-025-00214-z

3. Ssebibubbu S, Ssekamwa F, Muhumuza N, Mulumba M. Reforming Uganda’s digital health data systems: A policy analysis for inclusive, equitable, and decolonised data governance. Digital Health. 2026 Jan;12 10.1177/20552076251408532

4. Ministry of Health, Uganda. Strategic Health Plan 2020/21–2024/25. Kampala: Ministry of Health; 2021. https://library.health.go.ug/file-download/download/public/1524 [Last accessed: 11/03/2026]

5. National Planning Authority-Uganda. Third National development plan (NDPIII) 2020/21-2024/25. Kampala; 2020. https://budget.finance.go.ug/sites/default/files/NDPIII.pdf [last accessed: 11/03/2026]

6. National Planning Authority. Uganda Vision 2040: a transformative strategy for Ugandan society from a peasant to a modern, prosperous country within 30 years. Kampala; 2013. https://npa.go.ug/wp-content/uploads/2023/03/VISION-2040.pdf?x56883 [last accessed: 11/03/2026]

7. Ministry of Health Uganda. The Uganda Health Information and Digital Health Strategic Plan 2020/21–2024/25. Kampala: Ministry of Health; 2021 https://library.health.go.ug/file-download/download/public/1642 [last accessed 11/3/2026].

8. Huang F, Blaschke S, Lucas H. Beyond pilotitis: taking digital health interventions to the national level in China and Uganda. Globalization and health. 2017 Jul 31;13(1):49. DOI 10.1186/s12992-017-0275-z

9. Kiberu VM, Mars M, Scott RE. Barriers and opportunities to implementation of sustainable e-Health programmes in Uganda: A literature review. African Journal of Primary Health Care and Family Medicine. 2017 Feb 22;9(1):1–0. https://hdl.handle.net/10520/EJC-9762b8fd2

10. Namatovu HK, Semwanga AR, Kiberu VM, et al. Barriers and facilitators of eHealth adoption among healthcare providers in Uganda–a quantitative study. InInternational Conference on e-Infrastructure and e-Services for Developing Countries 2021 Dec 1 (pp. 234–251). Cham: Springer International Publishing. 10.1007/978-3-031-06374-9_15

11. Uganda Learning Hub for Immunisation Equity. Utility of Data Capture Platforms for Identifying Zero Dose Children in Uganda. Kampala: Ministry of Health; October 2024. Available from: https://www.unicef.org/uganda/media/6376/file/Zero%20Dose%20Data%20Platforms%20Report%202024.pdf [Last accessed: 12/03/2026]

12. United Nations Children’s Fund, Nutrition Information in Routine Reporting Systems: A Landscape Analysis for UNICEF’s Eastern and Southern Africa Region, UNICEF Eastern and Southern Africa Regional Office, 2020 [Link, last accessed 10/03/2026]

13. Government of Uganda, Office of the Prime Minister. Uganda Nutrition Action Plan II 2020/21–2024/25. Kampala: Office of the Prime Minister; 2020. https://scalingupnutrition.org/sites/default/files/2022-06/national-nutrition-plan-uganda.pdf [last accessed 10/3/2026].

14. World Health Organization, United Nations Children’s Fund (WHO & UNICEF). Strengthening national nutrition information systems: Annual gathering report of the EC-NIS project. Geneva: World Health Organization; 2022. Available from: https://cdn.who.int/media/docs/default-source/nutrition-and-food-safety/ec-nis-project/ec-nis-project-2022-annual-gathering-report.pdf [Last accessed 18/03/2026]

15. National Information Platform for Nutrition (NIPN). Uganda National Information Platform for Nutrition: Generating evidence for nutrition policy and decision-making. Kampala: Government of Uganda / NIPN Secretariat; 2024. https://www.nipn-nutrition-platforms.org/nipn-guidance-notes/

16. Uganda - Food Systems Dashboard

17. Ddamba A, Nsubuga B, Kamabare M, Abaho E, et al. Factors influencing the availability and use of electronic medical records systems in public health facilities in Uganda: a cross-sectional assessment. BMC Medical Informatics and Decision Making. 2025 Oct 10;25(1):372. 10.1186/s12911-025-03190-6

18. Kiwanuka SN, Kabwama SN, Namuhani N, et al. Barriers and facilitators of the effective use of DHIS2 data to improve program planning and monitoring in Uganda: a sequential mixed methods study. Oxford Open Digital Health. 2026 Jan 14 10.1093/oodh/oqag002

19. Alunyu Egwar A, Amiyo MR, Nabukenya J. Framework for standardizing digital health in resource-constrained settings: a case study of Uganda’s digital health communication infrastructure. Oxford Open Digital Health. 2024; 10.1093/oodh/oqae018

